# Discovering Novel Genome Mutations Associated with Liver and Pancreas Disorders: Insights from a Radiogenomics Study

**DOI:** 10.1101/2025.01.14.25320567

**Authors:** Mohamad Koohi-Moghdam, Varut Vardhanabhuti, Junwen Wang, Albert Chan, Kyongtae Tyler Bae

**Affiliations:** Department of Diagnostic Radiology, Li Ka Shing Faculty of Medicine, The University of Hong Kong, Pok Fu Lam, Hong Kong SAR, PR China; Division of Applied Oral Sciences and Community Dental Care, Faculty of Dentistry, The University of Hong Kong, Hong Kong SAR, PR China; Liver Transplant Center, Queen Mary Hospital, Department of Surgery, The University of Hong Kong, Hong Kong SAR, PR China

## Abstract

Radiogenomics provides a powerful approach to identify genetic variants linked to imaging-derived phenotypes, offering insights into the genetic underpinnings of organ function and disease susceptibility. The liver and pancreas are critical to metabolism, digestion, and detoxification, with their dysfunction leading to conditions such as pancreatitis, diabetes, and cancer. This study used a genome-wide association study (GWAS) to identify genetic variants associated with liver and pancreas radiomics phenotypes derived from magnetic resonance imaging (MRI). We conducted a cross-sectional study using data from 38,844 unique subjects in the UKBiobank, each with available MRI scans and genome sequences. We identified several novel single nucleotide polymorphisms (SNPs) associated with liver and pancreas characteristics. Notable findings include associations with rs1800562 (HFE gene), rs58542926 (TM6SF2 gene), rs738409 (PNPLA3 gene), and rs855791 (TMPRSS6 gene). These findings may contribute to our understanding of the genetic basis of organ function and could potentially help in future preventive healthcare.

## Introduction

The liver and pancreas are vital organs responsible for numerous physiological processes, including metabolism, digestion, and waste detoxification ^1-3^. Dysfunction in these organs can lead to severe conditions such as non-alcoholic fatty liver disease (NAFLD), pancreatitis, diabetes, and various cancers ^4,5^. Among the factors contributing to these disorders, abnormal iron metabolism, fat deposition, and changes in organ volume have emerged as significant risk factors ^6-9^. Understanding the genetic underpinnings of these traits could provide valuable insights into the pathogenesis of liver and pancreas-related diseases and offer potential avenues for personalized healthcare interventions.

Advances in radiogenomics, which integrate imaging data with genomic information, have opened new opportunities to explore the genetic basis of complex traits ^10,11^. Magnetic resonance imaging (MRI), in particular, offers a non-invasive method to measure radiomics traits such as iron levels, proton density fat fraction (PDFF), and organ volume in the liver and pancreas ^12,13^. These imaging-derived phenotypes provide high-resolution, quantitative data, enabling the identification of genetic variants associated with specific organ characteristics. Genome-wide association studies (GWAS) have proven to be a powerful tool for uncovering genetic variants linked to complex traits, offering a comprehensive approach to identifying single nucleotide polymorphisms (SNPs) associated with various phenotypes ^14,15^.

While previous GWAS have focused on traditional biomarkers and clinical outcomes, the integration of radiomics and genomics remains relatively underexplored. This study aims to bridge this gap by conducting a large-scale GWAS of MRI-derived radiomics traits in the liver and pancreas, leveraging data from the UKBiobank ^16^, a vast repository of genetic and imaging data. By identifying genetic variants associated with liver and pancreas iron levels, PDFF, and organ volume, this study seeks to uncover novel genomic insights that could inform our understanding of organ-specific pathophysiologies, with a particular focus on metabolic disorders and iron homeostasis.

Our study identified several significant genetic variants associated with MRI-derived traits in the liver and pancreas. These findings highlight the genetic complexity of liver and pancreas traits, offering insights into metabolic disorders and potential pathways for new treatment. Beyond clinical implications, such discoveries could potentially help our understanding of the molecular mechanisms driving organ dysfunction, thus aiding in the development of personalized medical strategies ^17,18^. By integrating radiogenomics data, we hope to provide new insights that could potentially inform future diagnostic and therapeutic strategies for liver and pancreas disorders.

## Materials and methods

### Study Population and Data Sources

This cross-sectional study utilized data from the UK Biobank, a large population-based cohort comprising over 500,000 participants aged between 40 and 69 years at recruitment ^19^. We extracted data for 38,844 unique participants who had undergone both abdominal magnetic resonance imaging (MRI) and whole-genome sequencing. The population cohort in this study was from the UK Biobank [Application Number 78730] which received ethical approval from the North West Multicentre Research Ethics Committee (REC reference: 11/NW/03820). All participants gave written informed consent before enrolment.

### MRI Data Acquisition and Processing

Abdominal MRI scans were performed using Siemens Aera 1.5 T scanner (Syngo MR D13) (Siemens, Erlangen, Germany) following standardized protocols detailed in the UK Biobank imaging documentation ^20^. Briefly, MRI sequences included the acquisition of liver and pancreas images to assess iron content, PDFF, and organ volumes. Processed MRI-derived radiomics features for liver and pancreas were obtained through the UK Biobank Research Analysis Platform (UK Biobank RAP, ukbiobank.dnanexus.com). The radiomics traits extracted for each participant included, liver iron content, liver PDFF, liver volume, pancreas iron content, pancreas PDFF and pancreas volume. These measurements were generated using standardized image analysis pipelines provided by UK Biobank, which utilized advanced algorithms for quantification of organ-specific parameters.

### Radiomics Trait Thresholding and Cohort Selection

We focused on six MRI-derived radiomics traits, including liver iron levels, liver PDFF, liver volume, pancreas iron levels, pancreas PDFF, and pancreas volume. For each radiomics trait, we calculated the mean and standard deviation (SD) across the entire dataset. Participants were categorized into case and control groups based on whether their measurements exceeded a threshold defined as the mean plus one SD (mean+SD) of the respective radiomics feature. Participants with values higher than this threshold were assigned to the case group, representing elevated levels of the trait, while those with values below the threshold were assigned to the control group. Subjects lacking data for any of the radiomics features were excluded from the respective analyses. The number of participants in each group for each trait shown in Table 1.

**Table 1.** Demographic characteristics of the study population for each radiomics trait.

| **Radiomics Trait** |  | **Control** | **Case** |
| --- | --- | --- | --- |
| **Liver iron level** | Sample Size | 27,460 | 2,898 |
|  | Age | 54±7 | 56±7 |
|  | Sex |  |  |
|  | Male | 12,968 (47%) | 1,677 (58%) |
|  | Female | 14,492 (53%) | 1,221 (42%) |
| **Liver PDFF** | Sample Size | 26,837 | 3,521 |
|  | Age | 54±7 | 54±7 |
|  | Sex |  |  |
|  | Male | 12,510 (47%) | 2,135 (61%) |
|  | Female | 14,327 (53%) | 1,386 (39%) |
| **Liver volume** | Sample Size | 33,031 | 5,903 |
|  | Age |  |  |
|  | Sex | 55±7 | 52±7 |
|  | Male | 14,332 (43%) | 4,425 (75%) |
|  | Female | 18,699 (57%) | 1,478 (25%) |
| **Pancreas iron level** | Sample Size | 25,394 | 4,070 |
|  | Age | 54±7 | 56±7 |
|  | Sex |  |  |
|  | Male | 13,226 (52%) | 1,047 (26%) |
|  | Female | 12,168 (48%) | 3,023 (74%) |
| **Pancreas PDFF** | Sample Size | 25,246 | 4,218 |
|  | Age | 56±7 | 57±7 |
|  | Sex |  |  |
|  | Male | 11,360 (46%) | 2,913 (31%) |
|  | Female | 13,886 (56%) | 1,305 (69%) |
| **Pancreas volume** | Sample Size | 31,919 | 5,695 |
|  | Age | 55±7 | 52±7 |
|  | Sex |  |  |
|  | Male | 14,148 (44%) | 3,996 (70%) |
|  | Female | 17,771 (56%) | 1,699 (30%) |

### Genotype Data Preparation and Genome-Wide Association Study

Whole-genome sequencing data for the participants were obtained from the UK Biobank, which provided high-quality genotypic data generated using standard protocols ^21^. Data extraction focused on the exome sequences with PLINK-formatted files provided by UK Biobank. We used a combination of Python packages, including Pandas, PySpark, and dxpy, to retrieve and process the genome data. For the GWAS, we used both PLINK ^22^ and REGENIE ^23^ to analyze the association between genetic variants and MRI-derived radiomics traits. The genomic data were filtered to include only the participants with available radiomics traits, and necessary quality control (QC) steps were applied to ensure data integrity. The QC process included the following steps ^24^: 1) Individual and SNP Missingness: We applied filters to remove individuals and SNPs with high missingness rates. Specifically, we used PLINK2 with the --geno 0.1 option to exclude SNPs with a missing call rate > 10%, and the --mind 0.1 option to exclude individuals with a missing genotype rate > 10%. 2) Sex Discrepancy: We addressed inconsistencies between assigned and genetic sex by filtering participants where the reported sex (field p31) matched their genetic sex (field p22001). Participants with discrepancies were excluded from the analysis. 3) Minor Allele Frequency (MAF): To ensure sufficient power for detecting associations, we filtered out rare variants with a minor allele frequency (MAF) < 0.01 using the --maf 0.01 option in PLINK2. Additionally, we used the --mac 100 option to exclude variants with a minor allele count below 100. 4) Hardy-Weinberg Equilibrium (HWE): SNPs deviating from Hardy-Weinberg equilibrium were filtered out using the threshold --hwe 1e-15 in PLINK2. This helped ensure that only variants in equilibrium were retained for the analysis. 5) Heterozygosity Rate: We monitored the heterozygosity rate to detect outliers that could suggest underlying genotyping errors or sample contamination. This was handled as part of the QC metrics in the UK Biobank data pipeline. 6) Relatedness: To remove participants with close genetic relatedness, we excluded individuals with a kinship coefficient > 0.125 (field p22021). This ensured that only unrelated individuals were included in the analysis, reducing potential biases from familial relationships. 7) Population Stratification: We limited the analysis to participants of White British ancestry (field p22006), which helped mitigate potential confounding due to population stratification. Further, we included the top 10 principal components (PCs) as covariates in the association models. This adjustment was done to further correct for subtle population structure within the White British subset. All analyses were performed on the UK Biobank Research Analysis Platform (RAP) using large-scale computing resources suitable for handling genomic and MRI data.

### Post-GWAS Analysis and Plotting

After running the GWAS, we applied post-processing steps to identify significant genetic associations. First, we merged the GWAS results across all chromosomes into a single comprehensive results file to streamline analysis. To isolate the most promising genetic variants, we set a genome-wide significance threshold of p < 5×10-7. Variants meeting or exceeding this threshold were considered statistically significant and were subjected to further analysis. For visualization, we used LocusZoom ^25^ to generate both Manhattan Plots and QQ plots. Additionally, LocusZoom facilitated fine-mapping of the associated regions, allowing us to pinpoint potential causal variants within complex loci and providing deeper insights into the genetic architecture influencing liver and pancreas radiomics phenotypes.

### Network and Enrichment Analysis

To further understand the biological significance of the identified SNPs, we conducted SNP enrichment analysis using the web-based tool snpXplorer ^26^. This tool allowed us to perform enrichment analysis and generate plots to visualize the enriched pathways and biological processes associated with the SNPs identified in our study. For tissue enrichment analysis, we utilized the web-based tool Enrichr ^27^. This platform enabled us to analyze the genes linked to the significant SNPs, providing insights into their expression in different human tissues. Besides, we used Enrichr-KG ^28^ to perform network analysis to find the connection between the SNPs and the reported traits and diseases.

## Results

### Genetic variants significantly associated with liver and pancreas MRI-derived traits

Our genome-wide association study identified multiple single nucleotide polymorphisms (SNPs) significantly associated with liver and pancreas traits derived from MRI phenotypes (Fig S1-S6). Several SNPs showed strong associations with liver iron levels. The most significant was rs1800562 (-log10 p= 60.437), located within the HFE gene on chromosome 6, which is known to be involved in iron metabolism. Other notable associations included rs13219787 near H2BC17 (-log10 p= 19.966), rs3130253 near MOG (-log10 p= 14.288),

rs3094093 near MDC1 and MDC1-AS1 (-log10 p= 11.195), rs855791 within TMPRSS6 on chromosome 22 (-log10 p= 9.805), rs2233952 near PSORS1C1 and PSORS1C2 (-log10 p= 8.992), and rs11752919 near ZSCAN23 (-log10 p= 6.356), all on chromosome 6.

For liver PDFF, the strongest association was observed with rs58542926 (-log10 p= 45.923) near TM6SF2 and AC138430.1 on chromosome 19. Additionally, rs738409 within PNPLA3 on chromosome 22 (-log10 p= 37.377), rs429358 near APOE on chromosome 19 (-log10 p= 8.298), and rs72785308 near TMC5 on chromosome 16 (-log10 p= 7.05) showed significant associations. For liver volume, the most significantly associated SNP was rs4665972 (-log10 p= 7.7), located near the SNX17 gene on chromosome 2. Regarding pancreas PDFF, the most significant SNP was rs1341982 (-log10 p= 7.131), located near CDKN2C on chromosome 1. For pancreas volume, the top associations were rs2287990 near CTRB1 on chromosome 16 (-log10 p= 9.976), rs77581903 near AGAP12P on chromosome 10 (-log10 p= 8.925), and rs3734626 near RSPO3 and AL356534.1 on chromosome 6 (-log10 p= 6.322). The complete results can be found in Table 2 and Fig 1A.

**Table 2.** Genetic variants significantly associated with liver and pancreas MRI-derived traits are presented in this table. The genome-wide association study identified several single nucleotide polymorphisms (SNPs) with strong associations to specific traits.

| Trait | Marker | rsID | Nearest gene(s) | $-\log_{10} (p)$ |
| --- | --- | --- | --- | --- |
| Liver Iron | 6: 26,092,913 G/A | rs1800562 | HFE | 60.437 |
| Liver Iron | 6: 27,893,892 G/A | rs13219787 | H2BC17 | 19.966 |
| Liver Iron | 6: 29,666,235 A/G | rs3130253 | MOG | 14.288 |
| Liver Iron | 6: 30,711,851 T/A | rs3094093 | MDC1, MDC1-AS1 | 11.195 |
| Liver Iron | 22: 37,066,896 A/G | rs855791 | TMPRSS6 | 9.805 |
| Liver Iron | 6: 31,138,114 A/G | rs2233952 | PSORS1C1, PSORS1C2 | 8.992 |
| Liver Iron | 6: 28,435,826 T/C | rs11752919 | ZSCAN23 | 6.356 |
| Liver PDFF | 19: 19,268,740 C/T | rs58542926 | AC138430.1, TM6SF2 | 45.923 |
| Liver PDFF | 22: 43,928,847 C/G | rs738409 | PNPLA3 | 37.377 |
| Liver PDFF | 19: 44,908,684 T/C | rs429358 | APOE | 8.298 |
| Liver PDFF | 16: 19,444,146 G/T | rs72785308 | TMC5 | 7.05 |
| Liver Volume | 2: 27,375,230 T/C | rs4665972 | SNX17 | 7.7 |
| Pancreases PDFF | 1: 50,977,769 C/T | rs1341982 | CDKN2C | 7.131 |
| Pancreases Volume | 16: 75,224,719 C/T | rs2287990 | CTRB1 | 9.976 |
| Pancreases Volume | 10: 48,010,528 C/T | rs77581903 | AGAP12P | 8.925 |
| Pancreases Volume | 6: 127,150,646 C/T | rs3734626 | AL356534.1, RSPO3 | 6.322 |

**Figure 1.**
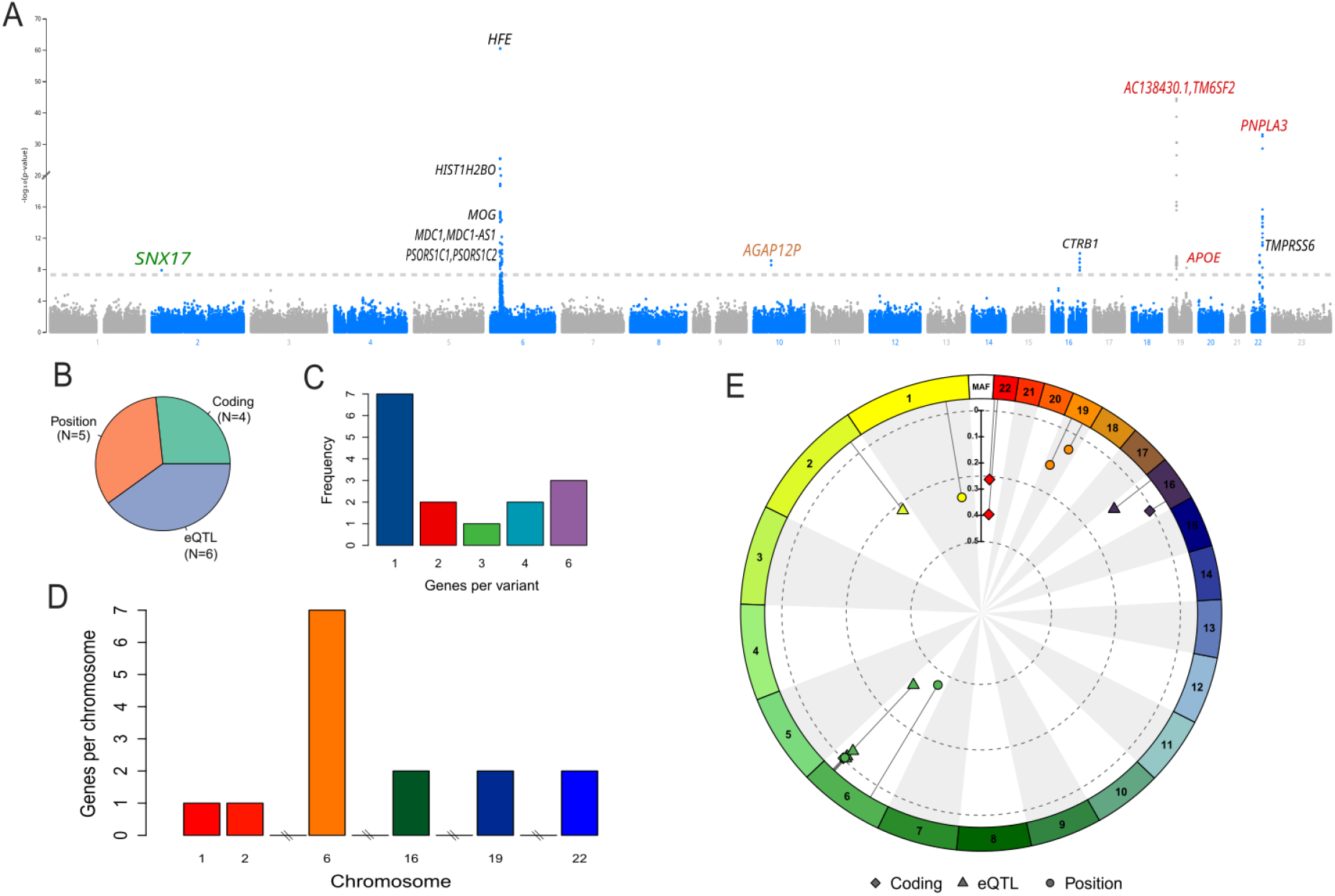
Overview of SNP associations with MRI traits. (A) Manhattan plot illustrating the results of GWAS. The x-axis represents genomic positions across chromosomes, while the y-axis shows the negative logarithm of -log10 (p-values) for each SNP. (B) Positional categories of identified SNPs, including five positional SNPs, four coding SNPs, and six eQTLs. (C) Number of genes affected by each SNP, highlighting pleiotropic effects where a single SNP influences multiple genes. (D) Chromosome-specific distribution of genes affected by SNPs, illustrating the higher density of functionally associated SNPs on certain chromosomes, such as chromosome 6. (E) Circular visualization of SNP allele frequencies across different genomic regions, including coding, regulatory, and non-coding regions.

### Genomic landscapes analysis highlight diversity of SNP associations with MRI traits

Our results show a variety of SNP types, categorized by their genomic locations and functions. Specifically, we identified five positional SNPs, four coding SNPs, and six eQTLs (expression Quantitative Trait Loci) (Fig 1B). To understand the impact of these SNPs on gene function, we analyzed the number of genes affected by each variant (Fig 1C). This analysis helps to highlight the pleiotropic effects of certain SNPs, demonstrating that a single variant can influence multiple genes. For instance, the variant rs4665972 on chromosome 2 is associated with several genes, including CAD, PPM1G, EIF2B4, GCKR, FNDC4, and KRTCAP3. Similarly, rs3130253 on chromosome 6 affects multiple HLA genes.

We also examined the distribution of genes affected by SNPs across different chromosomes (Fig 1D). Our findings indicate that certain chromosomes, such as chromosome 6, harbor a higher density of SNPs associated with multiple genes. This chromosome-specific analysis provides insights into the genetic architecture and potential hotspots for functional variants related to liver and pancreas disorder. The minor allele frequency (MAF) of the identified SNPs varies significantly, ranging from very low frequencies to more common variants. For example, rs72785308 on chromosome 16 has a MAF of 0.02596, while rs3734626 on chromosome 6 has a MAF of 0.4531. The sources of annotation for these SNPs are diverse, including coding regions, regulatory elements, and non-coding regions. This information is visually summarized in a circular chart, providing a comprehensive view of the allele frequencies and their functional annotations (Fig 1E).

### SNP enrichment analysis reveals key pathways in iron homeostasis

In addition to our initial SNP analysis, we conducted further investigations to understand the association of genes with specific traits. Our analysis revealed that certain genes are strongly associated with various reported traits (Fig 1A). Notably, the highest fraction of genes is associated with hemoglobin measurement, with nine genes implicated. Following closely, seven genes are linked to type II diabetes mellitus. Hematocrit and total cholesterol measurement each have five associated genes. Additionally, four genes are associated with aspartate aminotransferase measurement, liver fat measurement, liver disease biomarker, mean corpuscular hemoglobin concentration, reticulocyte count, and serum alanine aminotransferase measurement. This distribution highlights the significant role of these genes in various physiological and pathological processes. Importantly, the association with traits related to iron metabolism, such as hemoglobin measurement and liver disease biomarker, supports our hypothesis that these genes play critical roles in iron-related traits of MRI images.

To further understand the biological significance of the identified SNPs, we performed an enrichment analysis. The results show that several pathways are enriched with our SNPs (Fig 1B). The most significantly enriched pathway is “Hfe effect on hepcidin production” (WP:WP3924). This pathway is crucial for iron regulation and homeostasis. Additionally, other pathways such as “Iron metabolism disorders” (WP:WP5172) show moderate enrichment. These findings suggest that the identified SNPs play important roles in various biological processes, with a notable emphasis on iron metabolism.

**Figure 2.**
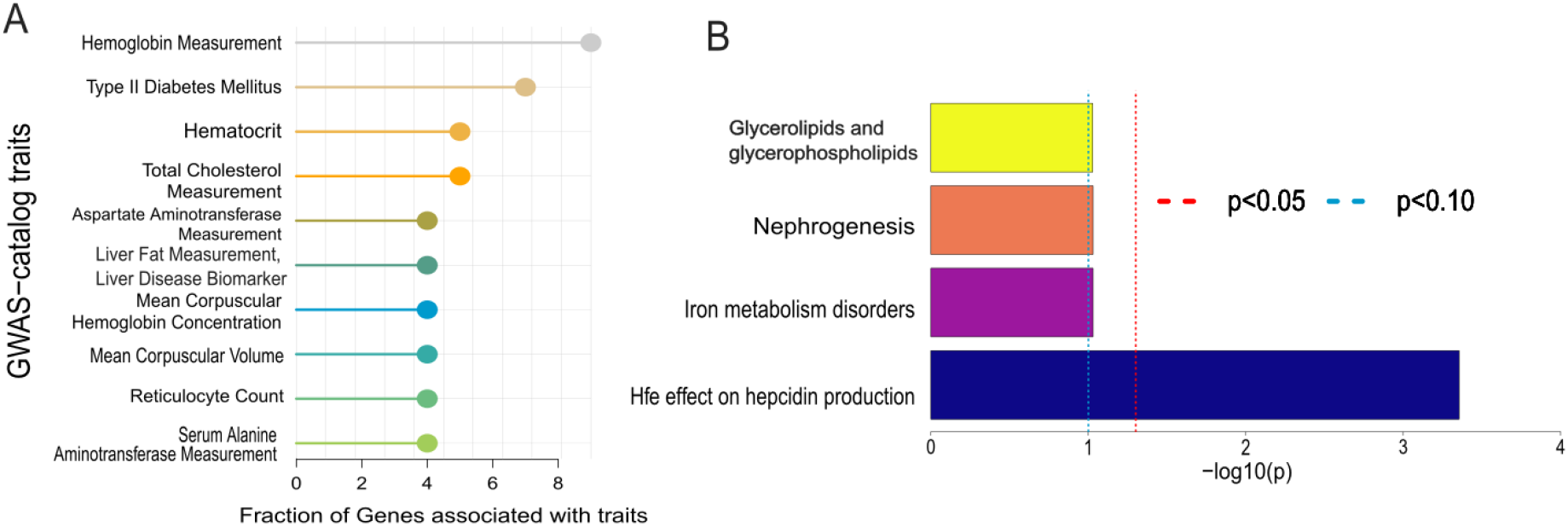
Gene enrichment and pathway enrichment analysis. (A) Gene-trait associations for various physiological and pathological processes, showing enrichment in traits related to iron metabolism, such as hemoglobin measurement and liver disease biomarkers. (B) Pathway enrichment analysis of SNPs, showing significant enrichment in pathways related to iron regulation, including “Hfe effect on hepcidin production” and “Iron metabolism disorders.

### Network analysis of SNP-associated genes shows connections to multiple traits and diseases

To further investigate the functional implications of the SNPs identified in our study, we conducted a network analysis to explore the connections between the genes associated with these SNPs and various diseases and GWAS traits. We used the GWAS catalog ^29^ and the Jensen Diseases database ^30^ for our network analysis (Fig 3A). Our analysis revealed significant associations between the identified genes and several traits and diseases. The top traits associated with the genes include hematology traits, iron status biomarkers, autism spectrum disorder or schizophrenia, hemoglobin concentration, and platelet count (Fig 3B). Additionally, several diseases were found to be significantly connected with the identified genes. These diseases include iron metabolism disease, fatty liver disease, peeling skin syndrome, hemochromatosis, and systemic scleroderma. The gene-trait and gene-disease network illustrates the complex interconnections between the identified genes and the various traits and diseases.

**Figure 2.**
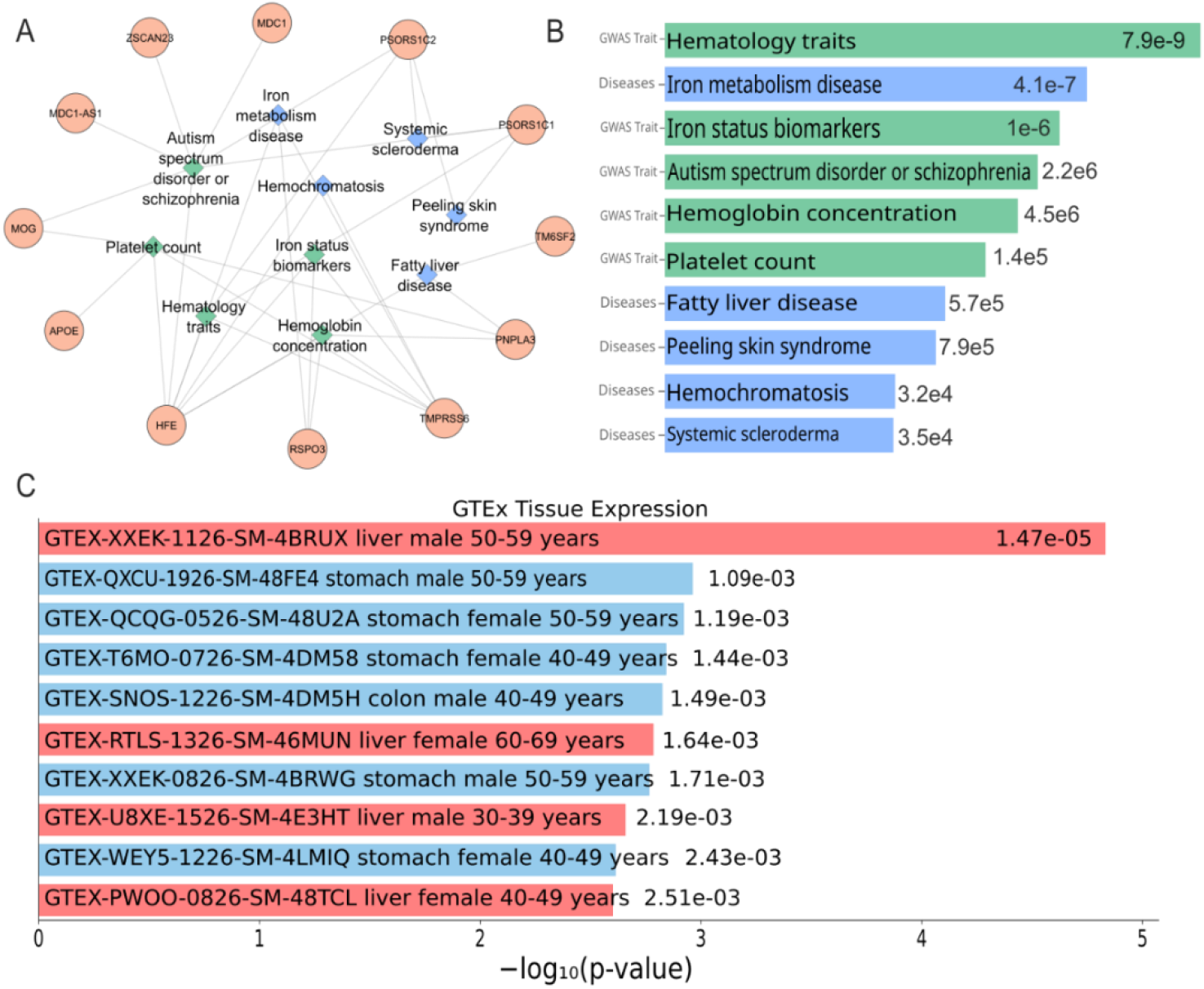
Network and tissue enrichment analysis of SNP-associated genes. (A) Network analysis of genes associated with identified SNPs, showing connections to various traits and diseases using data from the GWAS catalog and Jensen Diseases database. (B) Gene-disease network highlighting key diseases linked to the identified genes, including iron metabolism disease, fatty liver disease, peeling skin syndrome, hemochromatosis, and systemic scleroderma. (C) Tissue enrichment analysis of genes associated with SNPs, demonstrating significant expression in multiple tissues. The highest expression is observed in the liver across different demographic groups, particularly in males aged 50-59 years.

### Tissue enrichment analysis highlights liver as a key organ with high expression of genes associated with identified SNPs

To further understand the biological relevance of the tissue associated with the identified SNPs, we performed a tissue enrichment analysis. This analysis aimed to determine if these genes exhibit meaningful expression in specific tissues, particularly the liver and pancreas. We utilized the GTEx database ^31^ for this analysis. The results of our analysis revealed significant expression of these genes in various tissues and demographic groups (Fig 3C). Notably, the highest expression was observed in the liver of males aged 50-59 years, with a p-value of 1.47e-05. These genes also showed expression in the liver of females aged 60-69 years, with a p-value of 0.00164, suggesting that these genes are consistently expressed in liver tissue across different genders and age ranges. Additionally, gene expression was observed in the liver of males aged 30-39 years, with a p-value of 0.0025, further supporting their presence across various demographics.

Our findings also highlighted gene expression in the stomach across both genders and different age groups, although with slightly higher p-values, such as 0.0010 in males aged 50-59 years and 0.0014 in females aged 40-49 years. Similarly, the colon of males aged 40-49 years showed gene expression with a p-value of 0.0014. These results indicate a meaningful tissue-specific expression pattern, particularly in the liver, which could have implications for understanding biological functions or disease mechanisms in these tissues.

## Discussion

In this study, we performed a comprehensive genome-wide association analysis integrating radiomics phenotypes derived from MRI imaging with genetic data from the UK Biobank. Our aim was to identify genetic variants associated with liver and pancreas iron levels, PDFF, and organ volume. The radiogenomic approach allowed us to uncover several novel and known SNPs significantly associated with these traits.

### Genetic variants associated with liver iron levels reflect known iron metabolism pathways

Our GWAS identified multiple SNPs significantly associated with liver iron levels, most notably rs1800562 within the HFE gene on chromosome 6. This SNP exhibited the strongest association (-log10 p = 60.437) and is a well-established variant linked to hereditary hemochromatosis, a condition characterized by excessive iron accumulation in the body, particularly in the liver ^32,33^. The HFE gene plays a critical role in regulating iron absorption by modulating hepcidin production, a key hormone in iron homeostasis. Mutations in HFE disrupt this regulation, leading to iron overload.

Additionally, we identified rs855791 within the TMPRSS6 gene on chromosome 22 (-log10 p = 9.805). TMPRSS6 encodes matriptase-2, a protease that inhibits hepcidin expression. Variants in TMPRSS6 are known to affect iron metabolism by influencing hepcidin levels, thereby altering iron absorption and distribution ^34^. The identification of SNPs in HFE and TMPRSS6 highlights the pivotal role of the hepcidin pathway in liver iron regulation.

Other significant SNPs were located near genes such as H2BC17, MOG, MDC1, and ZSCAN23, all on chromosome 6. While these genes are not traditionally associated with iron metabolism, their proximity to the HLA region suggests potential immunological influences on iron homeostasis ^35^. Immune activation and inflammation can modulate hepcidin levels and iron distribution, indicating that these variants may have indirect effects on liver iron content ^36^.

Our SNP enrichment analysis further supports the involvement of iron metabolism pathways. The most enriched pathway was “HFE effect on hepcidin production” (WP:WP3924), confirming that the identified SNPs are functionally connected to critical mechanisms regulating iron levels in the liver. These findings reinforce the notion that genetic variants affecting iron metabolism pathways can significantly influence liver iron accumulation, which has implications for disorders such as hemochromatosis and iron-loading anemias.

### Associations with liver PDFF and volume reveal novel genetic factors in fat metabolism

For liver PDFF, which quantifies hepatic fat content, the strongest associations were observed with rs58542926 near the TM6SF2 gene (-log10 p = 45.923) and rs738409 within the PNPLA3 gene (-log10 p = 37.377). Both TM6SF2 and PNPLA3 have been previously implicated in non-alcoholic fatty liver disease (NAFLD) and steatohepatitis ^37,38^. Variants in these genes affect lipid metabolism in hepatocytes, influencing triglyceride accumulation and liver fat content.

The rs58542926 variant in TM6SF2 results in an amino acid change that impairs the secretion of very-low-density lipoprotein (VLDL), leading to intracellular lipid accumulation ^39^. Similarly, the rs738409 variant in PNPLA3 affects the protein’s lipase activity, promoting triglyceride retention in the liver ^40^. The identification of these SNPs verifies their role in hepatic steatosis and highlights their potential as genetic markers for NAFLD risk.

We also found associations with rs429358 near APOE and rs72785308 near TMC5. APOE is a key player in lipid transport and metabolism, with certain alleles linked to dyslipidemia and cardiovascular disease ^41^. Its association with liver fat content suggests a broader impact on lipid handling in the liver. rs72785308 in TMC5’s role is not yet fully understood, but its connection to the liver suggests it may play a part in how the liver processes fats. More research is needed to figure out its exact involvement.

Regarding liver volume, the most significant SNP was rs4665972 near the SNX17 gene (-log10 p = 7.7). SNX17 is involved in endosomal sorting and may influence receptor recycling and signaling pathways that affect hepatocyte proliferation and liver growth ^42^. This novel association points to potential genetic determinants of liver size, which could have implications for liver function and regenerative capacity.

### Pancreas-specific associations highlight genes involved in exocrine and endocrine functions

In the pancreas, we identified rs1341982 near CDKN2C (-log10 p = 7.131) associated with pancreas PDFF. CDKN2C is a cyclin-dependent kinase inhibitor involved in cell cycle regulation ^43^. Its role in pancreatic β-cell proliferation and function suggests that variants in CDKN2C may influence pancreatic fat deposition, potentially affecting insulin secretion and glucose homeostasis.

For pancreas volume, significant associations were found with rs2287990 near CTRB1 (-log10 p = 9.976), rs77581903 near AGAP12P, and rs3734626 near RSPO3. CTRB1 is a digestive enzyme precursor produced by pancreatic acinar cells. Variants near CTRB1 have been linked to type 2 diabetes and pancreatitis, suggesting a connection between exocrine function and pancreas size ^44^.

RSPO3 is involved in the Wnt signaling pathway, which plays a role in cellular proliferation and differentiation ^45^. Its association with pancreas volume may reflect its influence on pancreatic development and maintenance. These findings indicate that genetic factors affecting both exocrine and endocrine components contribute to pancreatic morphology and function.

### Network and tissue enrichment analyses emphasize the multifaceted roles of identified genes

Our network analysis revealed that the genes associated with the identified SNPs are connected to a variety of traits and diseases, particularly hematological traits and iron status biomarkers. The strongest associations were with hemoglobin measurement and type II diabetes mellitus, highlighting the interplay between iron metabolism, glucose regulation, and organ function.

The identified genes also showed significant expression in liver tissue across different age groups and genders, as indicated by our tissue enrichment analysis using Genotype-Tissue Expression (GTEx) data. The highest expression levels were observed in the liver of males aged 50-59 years (p = 1.47E-05), underscoring the liver-specific relevance of these genes. The expression patterns support the functional significance of these genes in hepatic physiology and their potential impact on liver-related disorders.

## Conclusion

By integrating MRI-derived radiomic phenotypes with genomic data, our study provides a nuanced understanding of how genetic variation influences organ-specific traits. The use of non-invasive imaging biomarkers allows for precise quantification of organ characteristics. This approach enhances the detection of genetic associations that may be missed when relying solely on traditional clinical phenotypes. Our findings highlight the value of radiogenomics in uncovering the genetic determinants of complex traits. The identified SNPs not only confirm known associations but also reveal novel variants that contribute to the variability in liver and pancreas characteristics. This integrative methodology paves the way for more personalized risk assessment and targeted interventions based on an individual’s genetic profile.

### Limitations and future directions

While our study provides valuable insights, there are limitations to consider. The cross-sectional design can’t reveal causality or temporal relationships. Longitudinal studies are necessary to determine how these genetic variants affect organ traits over time and their role in disease progression. The study population was derived from the UK Biobank, which predominantly includes individuals of European descent. Therefore, the generalizability of the findings to other ethnicities may be limited. Future studies should include more diverse populations to explore potential differences in genetic associations across ethnic groups.

Functional validation of the identified SNPs is also needed to elucidate their precise biological effects. Experimental studies using cellular and animal models can help confirm the mechanisms by which these variants influence liver and pancreas physiology. Finally, integrating additional omics data, such as transcriptomics and proteomics, could provide a more comprehensive understanding of the pathways involved and identify potential therapeutic targets.

## Data availability statement

The data supporting the findings of the study are available to researchers upon approval of an application to the UK Biobank (https://www.ukbiobank.ac.uk/researchers/).

## References

1 Field, K. M., Dow, C. & Michael, M. Part I: Liver function in oncology: biochemistry and beyond. The lancet oncology 9, 1092–1101 (2008).

2 Berasain, C., Arechederra, M., Argemí, J., Fernández-Barrena, M. G. & Avila, M. A. Loss of liver function in chronic liver disease: An identity crisis. Journal of hepatology 78, 401–414 (2023).

3 Karpińska, M. & Czauderna, M. Pancreas—its functions, disorders, and physiological impact on the mammals’ organism. Frontiers in physiology 13, 807632 (2022).

4 Pafili, K. & Roden, M. Nonalcoholic fatty liver disease (NAFLD) from pathogenesis to treatment concepts in humans. Molecular Metabolism 50, 101122 (2021).

5 Mederos, M. A., Reber, H. A. & Girgis, M. D. Acute pancreatitis: a review. JAMA 325, 382–390 (2021).

6 Kimita, W. & Petrov, M. S. Iron metabolism and the exocrine pancreas. Clinica Chimica Acta 511, 167–176 (2020).

7 Dongiovanni, P., Fracanzani, A. L., Fargion, S. & Valenti, L. Iron in fatty liver and in the metabolic syndrome: a promising therapeutic target. Journal of hepatology 55, 920–932 (2011).

8 Schmucker, D. L. Age-related changes in liver structure and function: Implications for disease? Experimental gerontology 40, 650–659 (2005).

9 Geng, Y., Faber, K. N., de Meijer, V. E., Blokzijl, H. & Moshage, H. How does hepatic lipid accumulation lead to lipotoxicity in non-alcoholic fatty liver disease? Hepatology international 15, 21–35 (2021).

10 Mazurowski, M. A. Radiogenomics: what it is and why it is important. Journal of the American College of Radiology 12, 862–866 (2015).

11 Liu, Z. et al. Radiogenomics: a key component of precision cancer medicine. British Journal of Cancer 129, 741–753 (2023).

12 Reeder, S. B. et al. Quantification of liver iron overload with MRI: review and guidelines from the ESGAR and SAR. Radiology 307, e221856 (2023).

13 França, M. & Carvalho, J. G. MR imaging assessment and quantification of liver iron. Abdominal radiology 45, 3400–3412 (2020).

14 Uffelmann, E. et al. Genome-wide association studies. Nature Reviews Methods Primers 1, 59 (2021).

15 Persyn, E. et al. Genome-wide association study of MRI markers of cerebral small vessel disease in 42,310 participants. Nature communications 11, 2175 (2020).

16 Collins, R. What makes UK Biobank special? The Lancet 379, 1173–1174 (2012).

17 Karlsen, T. H., Lammert, F. & Thompson, R. J. Genetics of liver disease: from pathophysiology to clinical practice. Journal of Hepatology 62, S6–S14 (2015).

18 Braat, H., Bruno, M., Kuipers, E. J. & Peppelenbosch, M. P. Pancreatic cancer: promise for personalised medicine? Cancer letters 318, 1–8 (2012).

19 UKBiobank. UKBiobank Data Showcase, <https://www.ukbiobank.ac.uk/enable-your-research/about-our-data> (2024).

20 UKBiobank. UKBiobank Body (abdominal) MRI scan, Version 1.0, <https://biobank.ndph.ox.ac.uk/crystal/ukb/docs/body_mri_explan.pdf> (2015).

21 Elliott, L. T. et al. Genome-wide association studies of brain imaging phenotypes in UK Biobank. Nature 562, 210–216 (2018).

22 Purcell, S. et al. PLINK: a tool set for whole-genome association and population-based linkage analyses. The American journal of human genetics 81, 559–575 (2007).

23 Mbatchou, J. et al. Computationally efficient whole-genome regression for quantitative and binary traits. Nature genetics 53, 1097–1103 (2021).

24 Marees, A. T. et al. A tutorial on conducting genome-wide association studies: Quality control and statistical analysis. International journal of methods in psychiatric research 27, e1608 (2018).

25 Pruim, R. J. et al. LocusZoom: regional visualization of genome-wide association scan results. Bioinformatics 26, 2336–2337 (2010).

26 Tesi, N., Van Der Lee, S., Hulsman, M., Holstege, H. & Reinders, M. J. snpXplorer: a web application to explore human SNP-associations and annotate SNP-sets. Nucleic acids research 49, W603–W612 (2021).

27 Kuleshov, M. V. et al. Enrichr: a comprehensive gene set enrichment analysis web server 2016 update. Nucleic acids research 44, W90–W97 (2016).

28 Evangelista, J. E. et al. Enrichr-KG: bridging enrichment analysis across multiple libraries. Nucleic acids research 51, W168–W179 (2023).

29 MacArthur, J. et al. The new NHGRI-EBI Catalog of published genome-wide association studies (GWAS Catalog). Nucleic acids research 45, D896–D901 (2017).

30 Pletscher-Frankild, S., Pallejà, A., Tsafou, K., Binder, J. X. & Jensen, L. J. DISEASES: Text mining and data integration of disease–gene associations. Methods 74, 83–89 (2015).

31 Carithers, L. J. & Moore, H. M. The genotype-tissue expression (GTEx) project. Biopreservation and biobanking 13, 307 (2015).

32 Katsarou, M. S. et al. Population-based analysis of the frequency of HFE gene polymorphisms: Correlation with the susceptibility to develop hereditary hemochromatosis. Molecular Medicine Reports 14, 630–636 (2016).

33 Katsarou, M.-S., Papasavva, M., Latsi, R. & Drakoulis, N. Hemochromatosis: hereditary hemochromatosis and HFE gene. Vitamins and hormones 110, 201–222 (2019).

34 Nai, A. et al. TMPRSS6 rs855791 modulates hepcidin transcription in vitro and serum hepcidin levels in normal individuals. Blood, The Journal of the American Society of Hematology 118, 4459–4462 (2011).

35 Zaimoku, Y. et al. HLA associations, somatic loss of HLA expression, and clinical outcomes in immune aplastic anemia. Blood, The Journal of the American Society of Hematology 138, 2799–2809 (2021).

36 Crux, N. B. & Elahi, S. Human leukocyte antigen (HLA) and immune regulation: how do classical and non-classical HLA alleles modulate immune response to human immunodeficiency virus and hepatitis C virus infections? Frontiers in immunology 8, 832 (2017).

37 Luukkonen, P. K. et al. Impaired hepatic lipid synthesis from polyunsaturated fatty acids in TM6SF2 E167K variant carriers with NAFLD. Journal of hepatology 67, 128–136 (2017).

38 Trépo, E., Romeo, S., Zucman-Rossi, J. & Nahon, P. PNPLA3 gene in liver diseases. Journal of hepatology 65, 399–412 (2016).

39 Musso, G. et al. TM6SF2 rs58542926 variant affects postprandial lipoprotein metabolism and glucose homeostasis in NAFLD [S]. Journal of lipid research 58, 1221–1229 (2017).

40 Johnson, S. M. et al. PNPLA3 is a triglyceride lipase that mobilizes polyunsaturated fatty acids to facilitate hepatic secretion of large-sized very low-density lipoprotein. Nature Communications 15, 4847 (2024).

41 Mahley, R. W. & Rall Jr, S. C. Apolipoprotein E: far more than a lipid transport protein. Annual review of genomics and human genetics 1, 507–537 (2000).

42 Cullen, P. J. Endosomal sorting and signalling: an emerging role for sorting nexins. Nature reviews Molecular cell biology 9, 574–582 (2008).

43 Li, G.-S. et al. Clinical significance of cyclin-dependent kinase inhibitor 2C expression in cancers: from small cell lung carcinoma to pan-cancers. BMC Pulmonary Medicine 22, 246 (2022).

44 Yazdanpanah, N. et al. Clinically relevant circulating protein biomarkers for type 1 diabetes: evidence from a two-sample Mendelian randomization study. Diabetes care 45, 169–177 (2022).

45 Chen, Z. et al. RSPO3 promotes the aggressiveness of bladder cancer via Wnt/β-catenin and Hedgehog signaling pathways. Carcinogenesis 40, 360–369 (2019).

